# The gut microbiome–amino acid pathway to emotional lability: a multi-step Mendelian randomization study

**DOI:** 10.64898/2026.10.06.26364911

**Authors:** Tao Jiang, Huabing Yang, Haofeng Li, Linling Li, Rui Wang, Yalin Zhang, Chen Zhong

## Abstract

Emotional lability (EL) is a prevalent cross-diagnostic symptom with unclear etiology. Given the potential role of the gut-brain axis in linking gut microbiota to EL, we adopted a multi-step Mendelian randomization (MR) framework to investigate the causal effects of gut microbiota on EL and the mediating role of blood metabolites. Bidirectional two-sample MR was first implemented to determine the causal relationship and direction between gut microbiota and EL, followed by multivariable MR (MVMR) to assess the direct contributions of individual microbial taxa. We then applied a multistep MR approach to identify gut microbiota specifically associated with EL-related psychopathology. Finally, the mediation effects of blood metabolites were quantified via the coefficient of variation method, and pathway analysis was conducted to verify the biological plausibility of the enriched metabolic pathways. Bidirectional MR detected 16 gut microbiota species associated with EL risk. MVMR identified three causally associated bacteria: RUG013 sp001486445 exerted the strongest positive effect (odds ratio [OR] = 1.570, 95% confidence interval [CI]: 1.090–2.261, P = 0.015), followed by CAG-485 sp002362485 (OR = 1.323, 95% CI: 1.080–1.619, P = 0.006). Each standard deviation (SD) increment in CAG-485 sp002362485 was linked to a 32.3% higher risk of EL. This bacterium was specific to EL and correlated with glycine/serine/threonine metabolites. Reverse MR analyses ruled out reverse causality.

**Author summary:** Emotional lability—defined as frequent, rapid shifts in mood and emotional state—is a pervasive symptom across many psychiatric disorders, and it is associated with poorer treatment outcomes and higher risks of self-harm. Despite its clinical significance, the biological causes of emotional lability remain largely unknown. Accumulating evidence points to a link between the gut microbiota (the community of microbes living in the human digestive tract) and brain function via the gut-brain axis, but it has remained unclear whether gut microbes directly cause changes in emotional stability, and what biological pathways might mediate such effects.

To answer these questions, we employed a multi-step Mendelian randomization approach. This method uses natural genetic variation as a tool to help distinguish causal relationships from simple correlations, reducing bias from confounding factors. We first tested the causal link between gut microbiota and emotional lability, then identified microbial markers specific to emotional lability by comparing them with other common psychiatric disorders. Finally, we investigated whether blood metabolites act as intermediate messengers that connect gut microbial activity to emotional regulation.

We identified 16 gut bacterial species that causally influence susceptibility to emotional lability, with three showing independent effects: two bacteria increase risk, while one is protective. The strongest risk-associated bacterium, CAG-485 sp002362485, was found to act in part by disrupting amino acid metabolism, particularly pathways involving glycine, serine, and threonine. We also confirmed that the effect flows from gut microbiota to emotional state, rather than emotional changes altering the gut microbiome.

These findings clarify the causal role of gut microbiota in emotional dysregulation and reveal key metabolic pathways that underpin the gut-brain connection. They not only improve our understanding of why emotional lability occurs, but also suggest new potential strategies for intervention, such as targeting gut microbes or related metabolic processes.

## Introduction

Emotional lability (EL) is a cross-diagnostic symptom characterized by rapid shifts in affect [1]. Its prevalence in the general population is estimated at up to 14%, with higher rates among adolescents and young women, and it is prevalent across multiple psychiatric disorders including depression, bipolar disorder, and schizophrenia [2, 3]. EL has been demonstrated to predict suicidal ideation and non-suicidal self-injury, and individuals exhibiting these behaviors may pose potential safety risks. In disorders such as Attention Deficit Hyperactivity Disorder (ADHD) and schizophrenia, heightened emotional instability is associated with increased aggression, which may complicate clinical management. Furthermore, EL correlates with poor outcomes [4, 5, 6]. Furthermore, research evidence indicates that emotional instability is a treatable clinical phenomenon, with pharmacotherapy representing a reasonable treatment option [7]. Thus, timely identification of EL symptoms in patients is crucial during clinical diagnosis and treatment [8]. However, current approaches to identifying, intervening, and screening for EL beyond the symptom dimensions of primary mental disorders remain inadequate. Understanding the etiology of EL offers valuable solutions and research directions. Thus, elucidating the causes of emotional dysregulation, enabling its precise identification, and facilitating early intervention are of paramount importance [7].

The gut microbiota is a complex ecological community that has evolved over thousands of years to form intricate relationships with its host [9]. The gut-brain axis (GBA), involving neural, immune, endocrine, and metabolic pathways, has been demonstrated to influence host mood and behavior [10]. Germinal animal studies indicate that the absence of gut microbiota can lead to severe behavioral abnormalities and neurodevelopmental deficits. Conversely, the administration of specific microbial strains can alter mood and cognition, further supporting the role of gut microbes in higher brain functions [11, 12]. This evidence suggests the gut microbiome plays a central role in the etiology of emotional instability. However, the relationship between gut microbiota and EL remains poorly understood. Unraveling these complex interactions holds promise for alleviating the substantial personal and societal burden of EL while opening new avenues for intervention [13].

Metabolites constitute essential elements within metabolic pathways, functioning as both substrates and terminal products that regulate vital cellular functions including energy production and storage, signal transduction, and apoptosis [14]. Substantial evidence implicates metabolic dysregulation as a pivotal contributor to the pathophysiology of psychiatric disorders, with documented abnormalities observed across conditions such as bipolar disorder and schizophrenia [15, 16]. Notably, emerging research underscores the role of the gut microbiota in modulating host metabolism, largely through the generation of bioactive metabolites capable of influencing cerebral function and behavior. Through the metabolism of dietary constituents, the gut microbiota yields a diverse array of metabolites, including short-chain fatty acids (SCFAs), tryptophan derivatives (e.g., serotonin and kynurenine pathway metabolites), and bile acids.

Metabolites such as bile acids can translocate across the intestinal epithelium into systemic circulation, thereby potentially exerting effects on the central nervous system [17, 18]. Nevertheless, significant questions persist regarding whether specific gut-derived metabolites are associated with mood instability and the mechanistic pathways that might underpin such associations remain elusive. Furthermore, while the gut-metabolic axis is increasingly implicated in diverse psychiatric conditions, its precise operational mechanisms are not fully delineated. This salient knowledge gap highlights the critical need to elucidate the pathways through which gut microbiota and their metabolic products may influence the pathogenesis of mood instability.

Mendelian randomization (MR) is a method for causal inference that uses genetic variants as instrumental variables to emulate randomization akin to randomized controlled trials [19, 20]. By leveraging genetic variants strongly associated with an exposure (e.g., gut microbiota), MR reduces confounding from environmental factors and minimizes reverse causation, thereby offering more reliable evidence on potential causal effects—such as those of gut microbiota on emotional instability [21].

## Materials and methods

### 2.1 MR design and data sources

As illustrated in Fig 1, this Mendelian randomization (MR) study was conducted in three phases, with the methodology adhering to the STROBE-MR reporting checklist (S1 Table) [22]. In Phase 1, we performed univariable MR analyses to investigate the causal association between gut microbiota and emotional lability (EL), and to eliminate reverse causality. Gut bacterial taxa with significant causal associations were subsequently subjected to multivariable MR analyses for further validation. In Phase 2, we investigated the associations of gut microbiota with psychiatric disorders frequently comorbid with EL; after excluding overlapping bacterial taxa, we sought to identify gut microbiota specifically associated with EL. In Phase 3, to elucidate the potential mechanisms by which the identified gut microbiota modulate EL, we first performed bidirectional MR analyses between serum metabolites and EL, followed by pathway enrichment analyses of the implicated metabolites. We then employed the difference-in-coefficients approach to quantify the overall genetically predicted effect of metabolite-associated gut microbiota on EL susceptibility.

**Fig 1.**
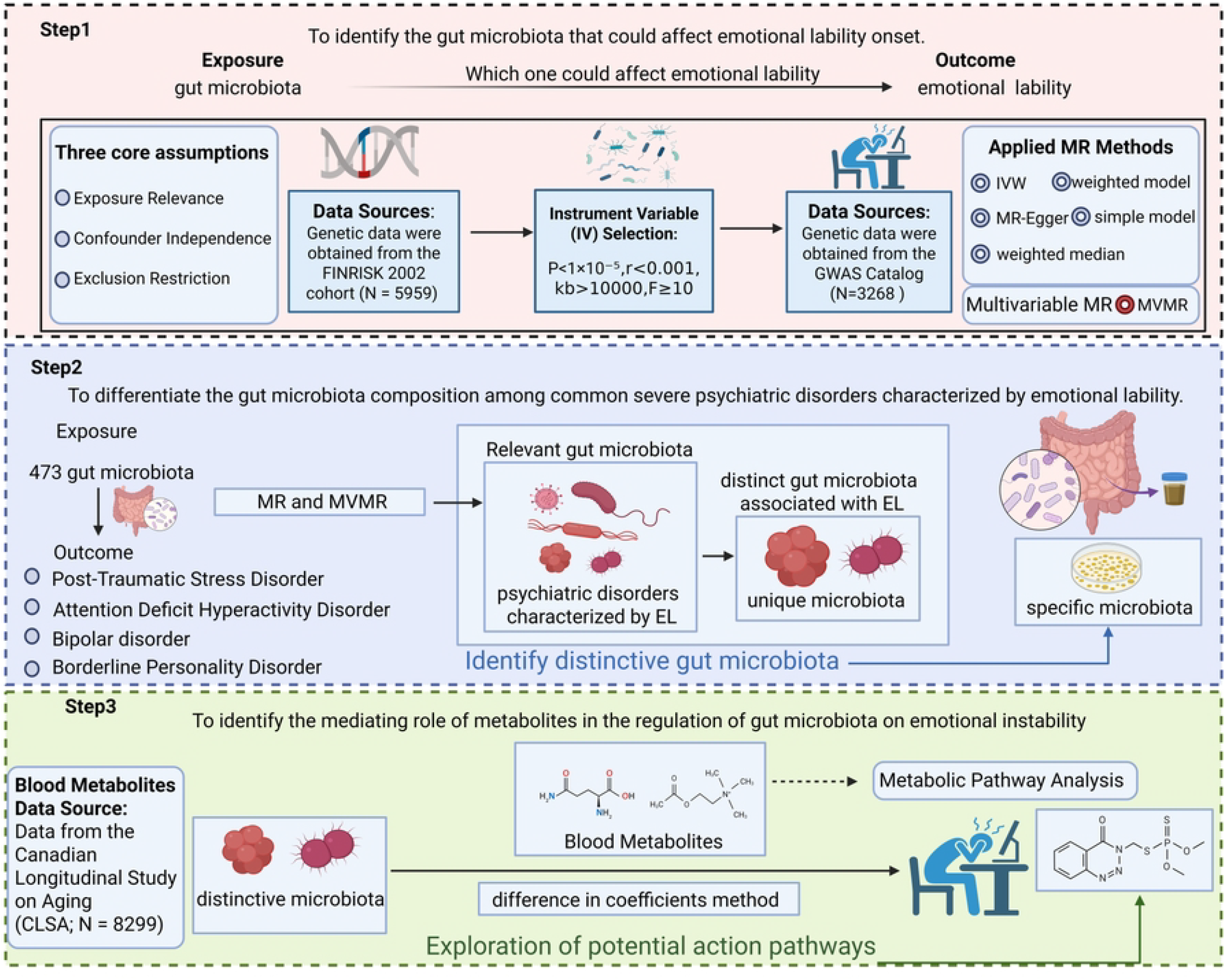
study design. Overview of the MR study design. EL, emotion lability; GWAS, genome-wide association study; MR, Mendelian randomization; IVW, inverse variance weighted; MVMR, multivariable MR.

Subsequent metabolic pathway enrichment analyses were conducted to compare EL-enriched metabolic pathways with those derived from the complete metabolite panel, aiming to identify gut microbiota-mediated core metabolic pathways underlying EL and clarify the regulatory role of gut microbiota in EL-associated metabolic processes. In mediation analyses, the total causal effect of gut microbiota on EL was defined as *β*_all_, the mediating effect as *β*_1_*β*_2_, the direct causal effect as *β*_all_ *− β*_1_*β*_2_, and the proportion of the total effect mediated as *β*_1_ *× β*_2_*/β*_all_.

### 2.2 Data sources

#### 2.2.1 Gut microbiota data

Gut microbiota-associated data are sourced from the GWAS catalog (https://www.ebi.ac.uk/gwas/). This catalog contains comprehensive aggregated data on genetic variations in the gut microbiome (GCST90032172 to GCST90032644) from 5,959 European individuals [23]. It includes 11 phyla, 18 classes, 24 orders, 62 families, 145 genera, and 213 species.

#### 2.2.2 Serum metabolites

Data from the Canadian Longitudinal Study on Aging (CLSA; *N* = 8299, European ancestry) were used for mediation analysis. Aggregate statistics are publicly available via the NHGRI-EBI GWAS Catalog (accessions GCST90199621–GCST90201020).

#### 2.2.3 Mood instability and other psychiatric disorders

- **EL:** The EL dataset from the GWAS catalog comprises 3,268 samples (dataset: GCST90013452), with participants completing a series of personality and health-related questionnaires. Using the original dataset by Amelang et al. and reanalyzing it with principal components and Varimax rotation via the R psychology library yielded identical results. Regression factor scores were calculated for the emotional instability personality dimension, with all participants being of European ancestry.
- **ADHD:** This data originates from the PGC database (https://pgc.unc.edu/). The PGC integrated dataset was meticulously compiled by Anders D. Børglum et al. in 2023 through a comprehensive analysis, establishing it as the most comprehensive ADHD GWAS dataset to date. This dataset integrates data from 13 cohorts, encompassing 38,691 ADHD patients and 186,843 controls—nearly doubling the sample size and making it one of the largest studies to date [24].
- **PTSD:** GWAS data for PTSD originate from the Psychiatric Genomics Consortium (PGC). Analyses encompass 60 distinct PTSD studies. Traumatic exposure includes civilian, military events, or both, typically occurring during childhood. Most controls comprise healthy, non-traumatized individuals. Additionally, we selected data from European ancestry, comprising 23,212 cases and 151,447 controls [25].
- **Bipolar disorder:** Data on bipolar disorder (BD) primarily originated from the Psychiatric Genomics Consortium (PGC). Specific diagnostic criteria are detailed on the official website of the Psychiatric Genomics Consortium (https://pgc.unc.edu/). Specifically, we utilized data from a recent meta-analysis on bipolar disorder bip2021 (ieu-b-5110).
- **Borderline personality disorder:** Summary data for BPD were derived from a genome-wide association study (GWAS) (https://gwas.mrcieu.ac.uk/datasets/finn-b-F5_EMOPER/), which included 2,637 BPD patients and 212,179 control subjects.

### 2.3 Instrumental variable assumptions and analysis

MR analysis requires the instrumental variable (IV) to satisfy three core assumptions: (1) the IV is strongly correlated with the exposure of interest, (2) the IV influences the outcome solely through the exposure (no horizontal pleiotropy), and (3) the IV is independent of any confounding factors. To ensure adherence to these assumptions, we satisfied them through the following approaches: (1) IV single nucleotide polymorphisms (SNPs) were initially selected using SNPs from exposure GWAS (*P <* 1 *×*10^*−*5^) as screening criteria; (2) SNPs in linkage disequilibrium (LD) (*r*^2^ *<* 0.001 and physical distance between any two SNPs *>* 10, 000 kb) were excluded. Subsequently, outcome GWAS data were extracted based on the filtered SNPs.

To assess potential weak instrumental bias, F-statistics were calculated. F-values *<* 10 indicated weak instruments; these were excluded to prevent bias and ensure the integrity of study findings. The F-statistic formula employed was as follows:

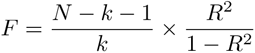

Where *N* denotes the sample size, *k* represents the number of instrumental variables employed, and *R*^2^ quantifies the extent to which the instrumental variables explain the exposure. *R*^2^ is calculated as follows:

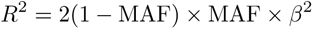

This study employed five distinct methods, each capable of yielding robust evidence under varying conditions. The inverse variance weighting (IVW) method, which proves most effective under the assumption of valid instrumental variables, served as the primary analysis. However, it may be susceptible to the addition or removal of genetic variants.

Sensitivity analyses were conducted to assess the robustness of MR results. Cochran’s Q test within the IVW method was utilised to examine heterogeneity among SNPs in the analysis. The MR-Egger method was employed to assess horizontal pleiotropy, with *P <* 0.05 indicating potential horizontal pleiotropy. The MR-PRESSO method was used to identify potential outliers or horizontal pleiotropy within the data. The aforementioned series of analyses were repeated to evaluate heterogeneity and pleiotropy. All statistical analyses were performed using R version 4.5.1.

### 2.4 Assessment of heterogeneity and horizontal pleiotropy

To address potential heterogeneity, which may lead to differing causal effect estimates across genotypes, both the MR-Egger and IVW methods incorporate heterogeneity tests. Subsequently, Cochran’s Q statistic was calculated to quantify heterogeneity among SNPs. Significant associations were identified based on IVW-derived *P* values *<* 0.05. The MR-Egger method was employed to assess horizontal pleiotropy, wherein a single gene or genetic variant influences multiple phenotypes or traits, thereby violating the second IV assumption.

## Results

### 3.1 Screening for gut microbiota associated with EL

The IVW method proves highly effective for inferring causal relationships between exposure and disease outcomes. Consequently, among 473 exposures analysed via MR, we identified 16 bacterial groups associated with EL using IVW. Furthermore, multiple Mendelian randomization analyses revealed that the OR values for most exposures and outcomes indicated a significant impact of gut microbiota on EL (Table 1 Fig 2).

**Table 1.**
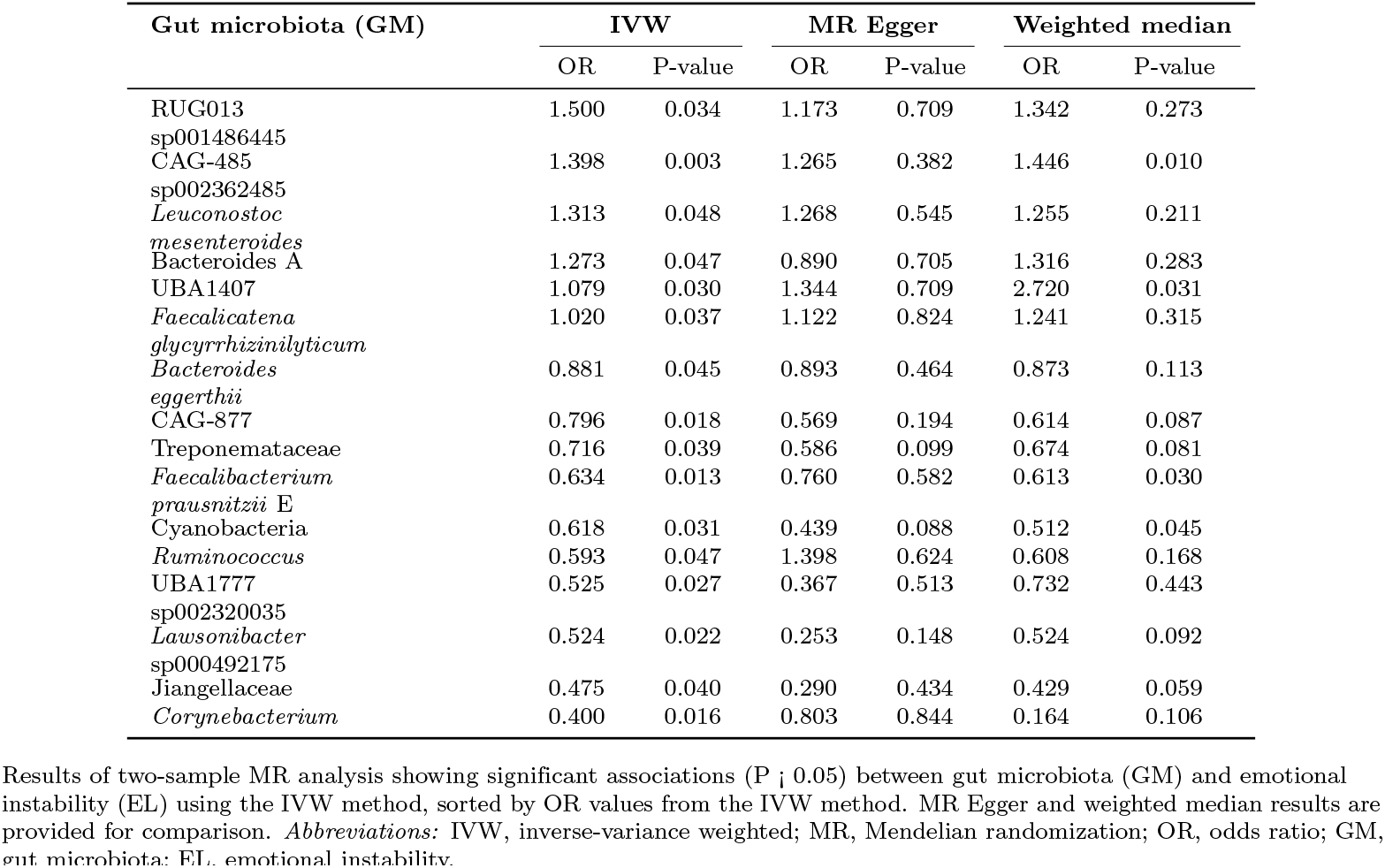
MR analysis of gut microbiota and emotional instability: significant IVW results with complementary methods.

**Fig 2.**
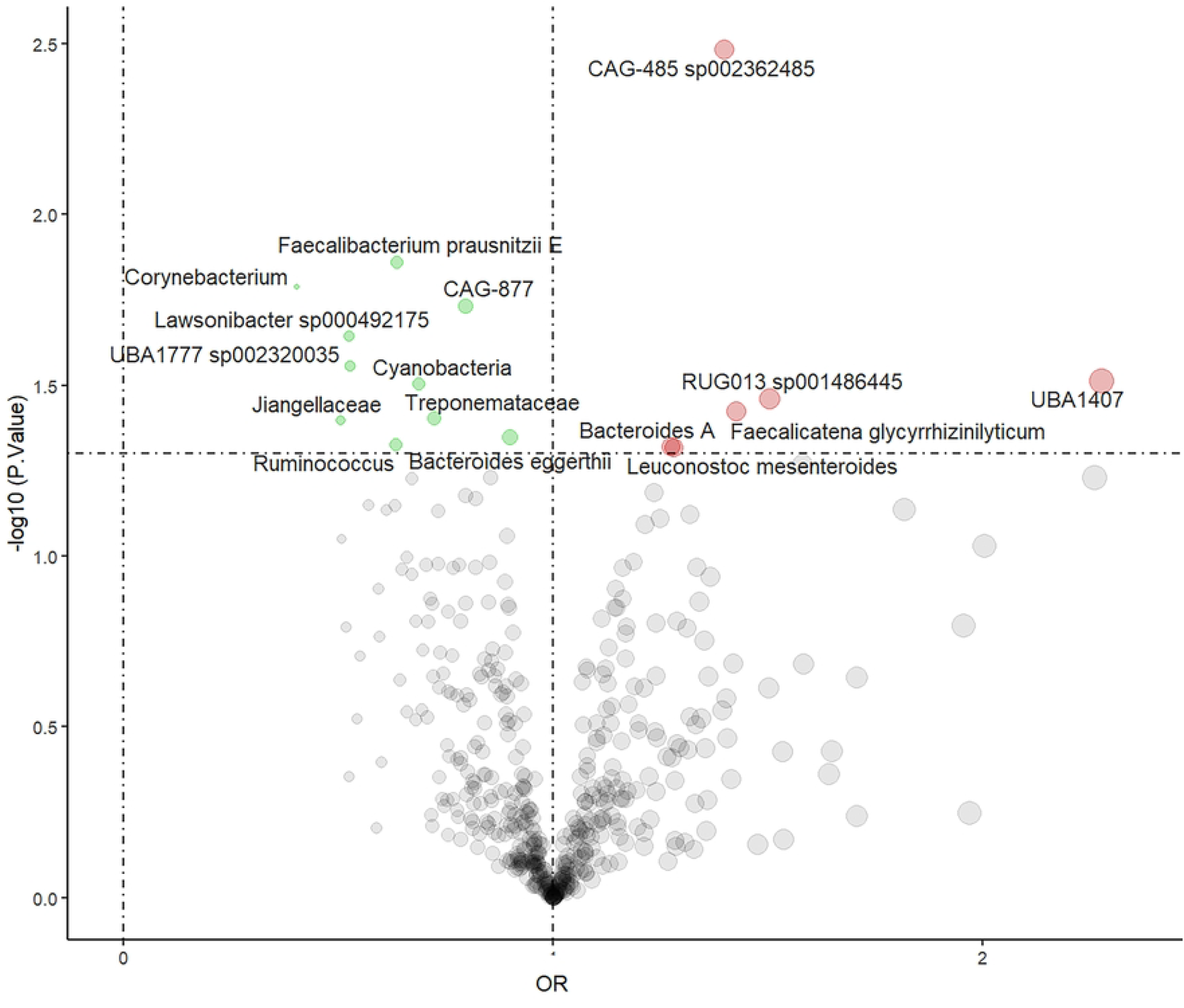
Volcano plot of gut microbiota after Mendelian randomization (MR) analysis.

The strongest positive effect was observed for RUG013 sp001486445: a one-standard-deviation increase in this bacterium was associated with a 50% higher risk of emotional instability (OR = 1.500, 95% CI: 1.029–2.187, *P* = 0.034). Second was CAG-485 sp002362485 (OR = 1.398, 95% CI: 1.118–1.749, *P* = 0.003), where a one-standard-deviation increase was associated with a 39.8% higher risk of emotional instability. Among protective factors, *Corynebacterium* exhibited the strongest effect.

Given the potential interrelationships identified among gut microbiota, we employed multivariable Mendelian randomization (MVMR) to calculate the direct effects of gut microbiota on EL, thereby identifying more specific microbiota (Table 2; Supplementary Table S2). Results indicated that CAG-485 sp002362485 (OR = 1.323, *P* = 0.006), RUG013 sp001486445 (OR = 1.570, *P* = 0.0153), and *Ruminococcus* (OR = 0.549, *P* = 0.0185) all exhibited statistical significance.

**Table 2.** Gut microbiota after multivariate Mendelian randomization (MVMR) analysis.

| Exposure | P-value | OR | SE | OR-CI |
| --- | --- | --- | --- | --- |
| CAG-485<br>sp002362485 | 0.006 | 1.323 | 0.103 | 1.080–1.619 |
| RUG013<br>sp001486445 | 0.0153 | 1.570 | 0.186 | 1.090–2.261 |
| *Ruminococcus* | 0.0185 | 0.549 | 0.254 | 0.334–0.904 |

### 3.2 Reverse two-sample MR analysis

In the reverse two-sample magnetic resonance analysis, we found that emotional instability exerted no significant causal influence on CAG-485 sp002362485, RUG013 sp001486445, or *Ruminococcus*. There was no overlap between EL-related instrumental variables (IV) and SNPs associated with these microbial taxa. Genetic instruments for EL indicated that the null causal estimates were robust against pleiotropy or shared genetic confounding.

### 3.3 Sensitivity analyses

Should horizontal pleiotropy exist, the IV may influence outcomes through pathways beyond the exposure factor, thereby violating the second MR assumption. We therefore calculated horizontal pleiotropy using the MR-Egger intercept, yielding results for CAG-485 sp002362485 (intercept = 0.010, *P* = 0.673), RUG013 sp001486445 (intercept = 0.013, *P* = 0.512), and *Ruminococcus* (intercept = *−*0.033, *P* = 0.192) all showed no horizontal pleiotropy.

Heterogeneity analysis revealed no evidence of heterogeneity for CAG-485 sp002362485, RUG013 sp001486445, or *Ruminococcus* in either the IVW (*P* = 0.929, 0.527, 0.438) or MR-Egger (*P* = 0.950, 0.574, 0.375) analyses.

MR-PRESSO identified outliers, with results indicating no significant outliers for CAG-485 sp002362485, RUG013 sp001486445, or *Ruminococcus*. Sensitivity analysis using the leave-one-out method confirmed the robustness of all reported associations, with no single SNP exhibiting disproportionate influence across the tested taxonomic units. To further validate the initial MR analysis results and ensure robustness (Fig. 3B), we applied stricter criteria requiring significance in at least two MR methods. Under these conditions, CAG-485 sp002362485 demonstrated statistically significant associations in the IVW and weighted median methods (*P <* 0.05). The MR-Egger estimate did not reach significance, potentially due to its limited power [26]. Notably, the majority of exposure-outcome relationships demonstrated good consistency across multiple Mendelian analyses (Fig. 3A).

**Fig 3.**
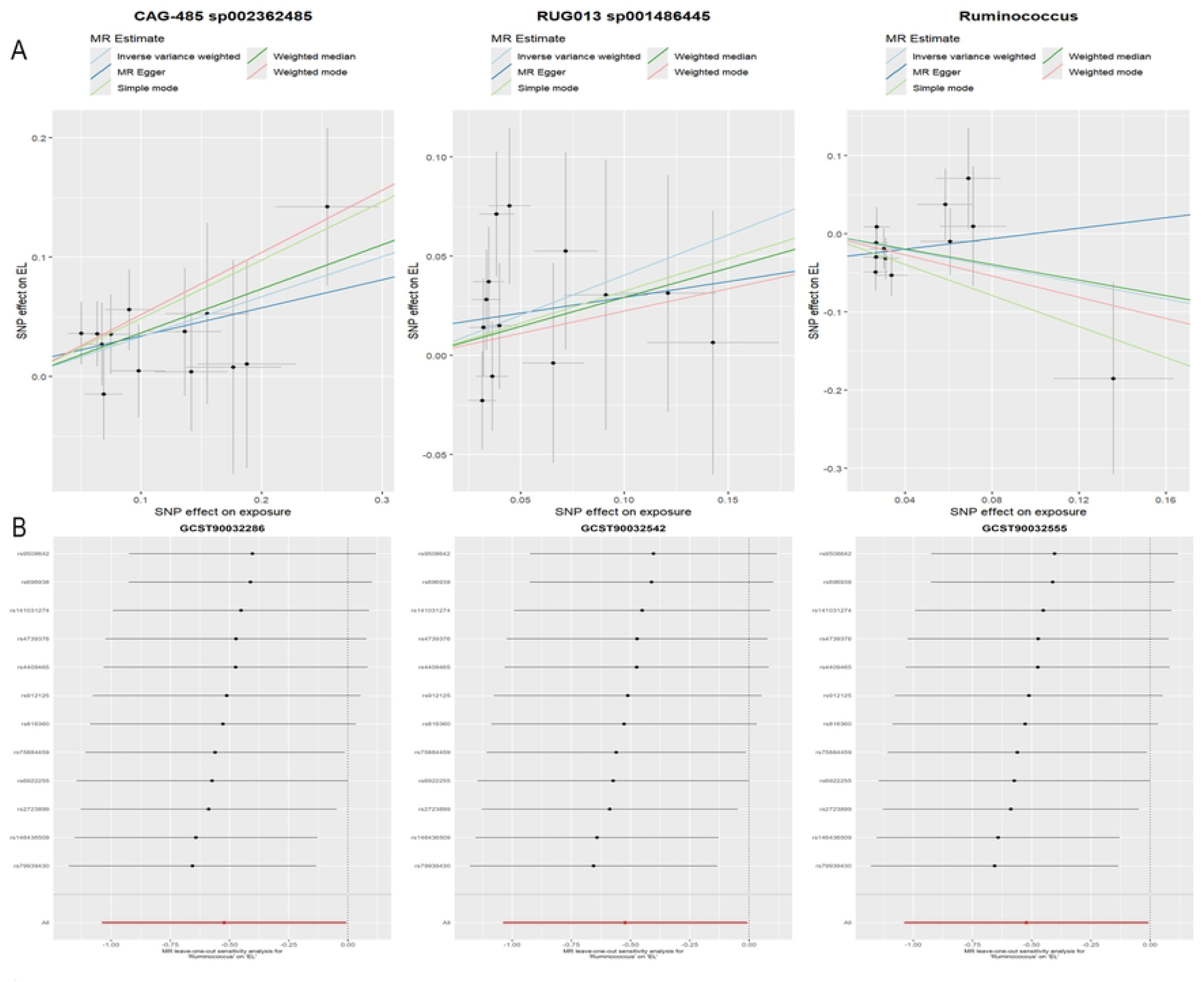
A. Scatter plot of three gut microbiota taxa against EL: CAG-485 sp002362485 and RUG013 sp001486445 show a positive association with EL risk, while Ruminococcus exhibits a protective effect; B. Leave-one-out sensitivity testing corresponding to the three gut microbiota taxa.

### 3.4 Screening for gut microbiota specific to EL

Subsequently, we applied two-sample and multivariate MR analyses to gut microbiota associated with common EL psychiatric disorders, employing the same screening framework to identify EL-specific gut microbiota. Two-sample Mendelian randomization results indicated that among 473 gut microbiota species, 31 were associated with ADHD, 30 with PTSD, 24 with bipolar disorder, and 21 with borderline personality disorder (Fig. 4A; Supplementary S3 Table). After MVMR, *Propionibacterium freudenreichii* (OR = 1.287, *P* = 0.020) was associated with ADHD, and *SAR324* (OR = 0.432, *P* = 0.003) with borderline personality disorder (Fig. 4B). However, after MVMR, gut microbiota previously associated with PTSD and bipolar disorder no longer showed significance (Supplementary Tables 4, 5).

**Fig 4.**
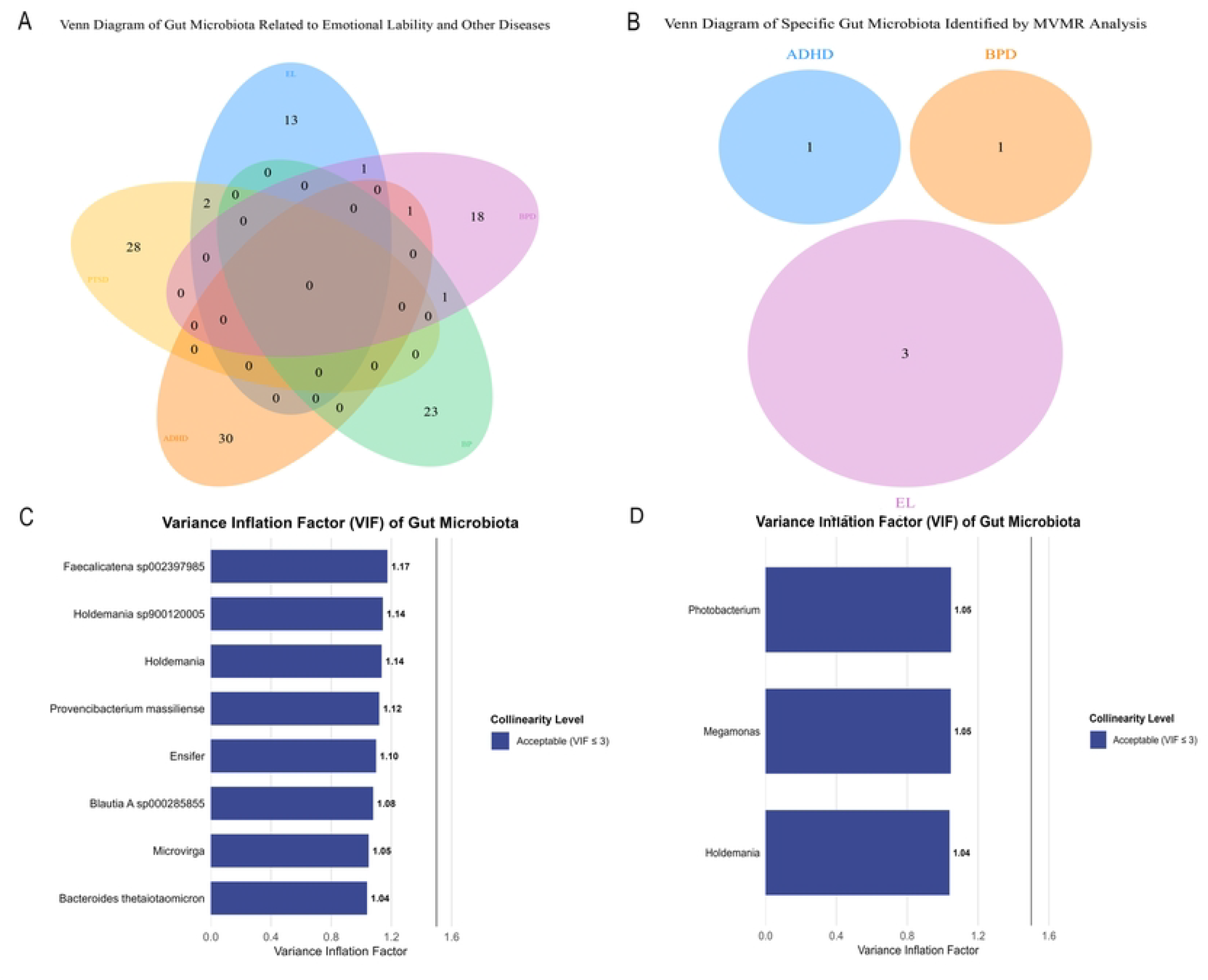
A. Venn diagram of factors associated with five mental disorders after screening via bidirectional Mendelian randomization (MR); B. Venn diagram of gut microbiota associated with three mental disorders after multivariate Mendelian randomization (MVMR) analysis; C. Bar chart of variance inflation factor (VIF) analysis for Post-Traumatic Stress Disorder (PTSD); D. Bar chart of variance inflation factor (VIF) analysis for bipolar disorder (BD).

Subsequently, we conducted variance inflation factor (VIF) analyses on gut microbiota with FDR ¡ 0.05 from the dual-sample Mendelian randomization screening to explore potential multicollinearity. Results demonstrated VIF values ¡ 5 and conditional number (*κ*) ¡ 10 across all three analyses (Fig. 4C, D). These findings support the conclusion that the identified gut microbiota possess independent genetic influences without multicollinearity [27, 28], suggesting their distinct yet synergistic alterations may correlate with disease risk.

### 3.5 Mediating effects

To investigate potential mechanisms underlying the relationship between GM and emotional instability, we analysed the mediating effects of 1,400 blood metabolites via multivariate Mendelian randomisation (MVMR) (Supplementary Table S6). Following multiple testing false discovery rate (FDR) correction (Benjamini–Hochberg, *q <* 0.05), we identified a set of statistically robust mediators for CAG-485 sp002362485. Among these, the strongest positive mediator was s-1-pyrroline-5-carboxylate (*β* = 0.271, OR = 1.311, CI: 1.161–1.642), mediating 15.52% of the effect, followed by N-lactoyl valine (*β* = 0.270, OR = 1.311, CI: 1.077–1.595) and cystine (*β* = 0.226, OR = 1.254, CI: 1.074–1.463), with mediating effects of 3.88% and 29.55% respectively (Table 3).

**Table 3.** Mediation analysis for EL.

| Exposure | Mediator | Outcome | Total effect | Mediator effect | Direct effect | Mediation effect proportion |
| --- | --- | --- | --- | --- | --- | --- |
| CAG-485 sp002362485 | s-1-pyrroline-5-carboxylate | EL | 0.335 | 0.052 | 0.283 | 15.52% |
| CAG-485 sp002362485 | n-lactoyl valine | EL | 0.335 | 0.013 | 0.322 | 3.88% |
| CAG-485 sp002362485 | cystine | EL | 0.335 | 0.099 | 0.236 | 29.55% |

### 3.6 Enrichment analysis

First, we performed two-sample Mendelian randomisation between 1,400 metabolites and emotional lability. After examining their directionality, sensitivity, heterogeneity, and horizontal pleiotropy, we identified 58 metabolites associated with EL. Formiminoglutamate showed the most significant association (*P* = 2.972*×*10^*−*4^, OR = 1.359), followed by orotate (*P* = 4.470*×*10^*−*4^, OR = 0.878). Following rigorous quality filtering and manual annotation (methods), enrichment analysis was conducted on the selected metabolites (Fig. 5A, C). Analysis of all metabolites revealed the top three metabolic pathways: histidine metabolism, glycine, serine and threonine metabolism, and arginine and putrescine metabolism.

Subsequently, enrichment analysis was performed on gut microbiota-associated metabolites using the same methodology. Metabolites linked to the gut microbiota showed significant enrichment in specific pathways (glycine, serine and threonine metabolism, *P* = 3.84 *×*10^*−*3^) (Fig. 5B, D). Notably, this pathway (glycine, serine, and threonine metabolism) also demonstrated high significance in the analysis of all metabolites, ranking as the second most enriched pathway (*P* = 4.18 *×* 10^*−*3^).

**Fig 5.**
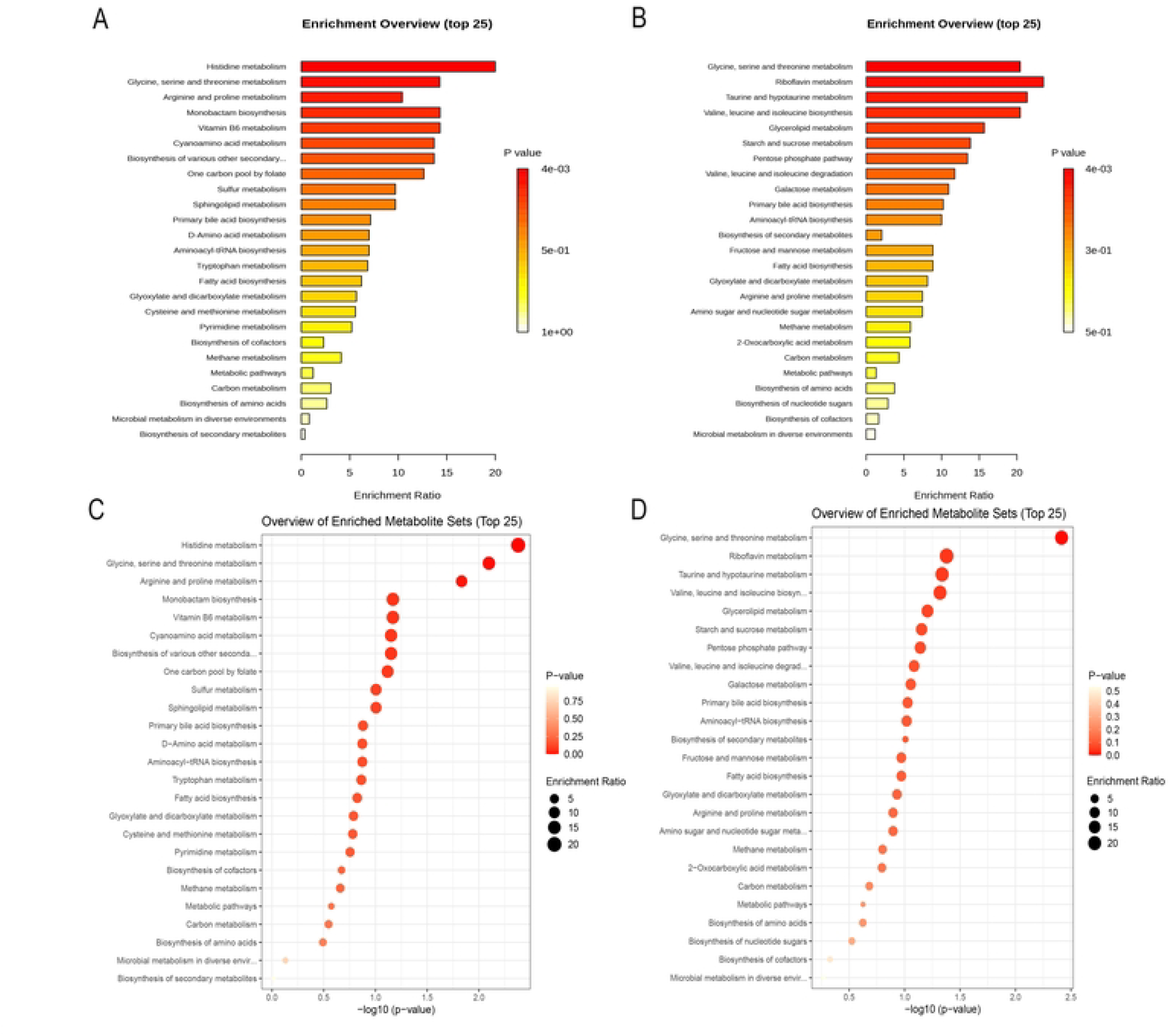
A. Bar chart of enriched metabolic pathways associated with emotional instability; B. Bar chart of enriched metabolic pathways mediating the effect of gut microbiota on emotional instability; C. Dot plot of enriched metabolic pathways associated with emotional instability; D. Dot plot of enriched metabolic pathways mediating the effect of gut microbiota on emotional instability.

## Discussion

Here, employing genetic variants as instrumental variables through Mendelian randomization analysis, we systematically investigated the role of gut microbiota in mood regulation. This study identified CAG-485 sp002362485 as a risk factor for mood instability. The effect magnitude (OR *≈*1.398) is comparable to that of established behavioral risk factors (e.g., smoking on cardiovascular disease), underscoring the biological relevance of this microbiota in mood dysregulation [29]. Consistent results across multiple MR methods, together with robustness confirmed by sensitivity analyses and heterogeneity tests, further support the specificity of this association. Mechanistic analyses indicate that the glycine, serine, and threonine metabolism pathways may mediate the influence of CAG-485 sp002362485 on emotional lability. Mediation analysis further revealed that gut microbiota play a key regulatory role in the metabolic modulation of affective instability. Collectively, these findings suggest that CAG-485 sp002362485 and its related metabolic pathways are more likely to represent potential pathogenic contributors to affective instability, rather than secondary microbial alterations resulting from emotional abnormalities. In summary, this study establishes a coherent link between specific gut microbiota, amino acid metabolism, and dysregulated emotional control. It not only provides new evidence for the multifunctional regulatory role of CAG-485 sp002362485 but also preliminarily delineates a “microbiota (CAG-485 sp002362485)–amino acid metabolism–brain function” regulatory network, offering novel directions for elucidating the microbial mechanisms underlying emotional disorders.

The human gut microbiota comprises trillions of microorganisms, and substantial evidence indicates that gut flora is closely associated with mental states, physical health, and psychological well-being [13]. The concept of the gut–brain axis (GBA) has evolved over decades, initially focused on neuroendocrine signaling but now widely recognized to include the gut microbiome as a key regulator of mood, cognition, and behavior [30]. Multiple studies have shown that increased abundance of Prevotellaceae is correlated with autoimmune phenomena [31]. CAG-485 sp002362485, as a specific member of an unannotated group within the Prevotellaceae family, exhibits significantly elevated abundance in experimental autoimmune uveitis (EAU) models, suggesting its potential link to inflammatory processes. This association may operate through a classical gut–brain pathway: “gut inflammation *→*activation of the hypothalamic–pituitary–adrenal (HPA) axis *→*neurotransmitter disruption” [32, 33]. Specifically, CAG-485 sp002362485 may secrete proteases capable of degrading the mucosal layer. These proteases can activate the TLR4 pathway, leading to downregulation of tight-junction proteins (e.g., ZO-1, claudin) in intestinal epithelial cells, thereby increasing intestinal permeability, exacerbating systemic inflammation, and ultimately contributing to neurotransmitter dysregulation and emotional disturbances [34, 35]. This perspective is further supported by observations of elevated Prevotellaceae abundance in chronic inflammatory conditions such as obesity [36].

Following the identification of gut-microbiota markers associated with mood instability through bidirectional and multivariable Mendelian randomization (MR) analysis, we further applied the same analytical framework to evaluate microbial markers for other psychiatric disorders in which mood instability represents a core clinical feature—including bipolar disorder, borderline personality disorder, attention-deficit/hyperactivity disorder, and post-traumatic stress disorder. Using consistent screening criteria, we compared microbiota linked to emotional instability with those associated with each of the above disorders, aiming to determine whether the identified markers robustly and specifically correlate with risk of emotional instability. The results indicated that *Propionibacterium freudenreichii* was associated with attention-deficit/hyperactivity disorder, whereas the SAR324 clade showed an association with borderline personality disorder. These observations suggest that gut-microbiota signatures predictive of distinct psychiatric disorders exhibit disease specificity, which aligns with existing clinical evidence and underscores their potential translational relevance [37, 38, 39]. In the analysis of PTSD and bipolar disorder using multivariable MR—which estimates the direct contribution of each microbial feature while accounting for the influence of other identified microbiota—we observed that alterations in these microbiota alone did not clearly distinguish their respective impacts on disease onset. Further evaluation using variance inflation factors confirmed that these microbiota contributed independent genetic signals and showed no evidence of multicollinearity, supporting the hypothesis that coordinated changes in gut-microbiota abundance may act synergistically to promote the development of the corresponding disorders.

Previous studies have proposed potential mechanisms underlying the causal relationship between gut microbiota and psychophysiological processes [40, 41, 42]. The mechanisms through which gut microbiota influence mental health are highly complex, involving neural, endocrine, immune, and metabolic pathways that connect the gut and the brain[43, 44]. Existing research confirms that gut microbiota can produce neuroactive substances by regulating amino acid metabolism or can influence host amino acid homeostasis, thereby modulating the synthesis and signaling of central neurotransmitters and subsequently affecting emotion-related behaviors [45]. The present findings suggest that gut microbiota may disrupt host neuro-metabolic homeostasis by reshaping metabolic flux through these pathways, ultimately contributing to emotional instability. The three metabolite mediators identified in this study—s-1-pyrroline-5-carboxylate, N-lactoyl-valine, and cystine—provide novel metabolite references linking metabolism to mental health. s-1-Pyrroline-5-carboxylate and 1-pyrroline-5-carboxylate (P5C) represent variant nomenclatures for the same compound. P5C serves as a key intermediate in proline and glutamate metabolism and is closely linked to glycine, serine, and threonine pathways. It is converted into glutamate semialdehyde via the phosphoglycerate pathway and subsequently cyclizes to form P5C. Insufficient serine supply reduces P5C synthesis, leading to decreased proline levels. As an osmotic buffer and synaptic transmission regulator in the central nervous system, proline deficiency can result in neuronal osmotic imbalance and synaptic dysfunction, thereby inducing emotional abnormalities such as anxiety and irritability [46]. N-lactoylated amino acids (Lac-AA) are emerging as important metabolites with diverse physiological functions. Prior research has demonstrated their notable role in certain gut bacterial functions and their association with gut microbial dysbiosis. Lactoyl-valine (Lac-Val) also shows significant correlations with clinical parameters including white blood cell count and glucose levels, and is enriched in inflammatory pathways such as NF-*κ*B and TNF. This forms a closed-loop mechanism that aligns with the exacerbated inflammation observed alongside elevated levels of CAG-485 sp002362485[47, 48]. Cystine, a dimer of cysteine, requires serine as an essential precursor for its biosynthesis. Serine undergoes a sulfur-transfer reaction catalyzed by serine acetyltransferase and cystathionine *β*-synthase to form cysteine, which subsequently dimerizes into cystine. Abnormal serine metabolism directly impairs cystine synthesis. Previous studies have reported that cystine represents a terminal metabolite of homocysteine metabolism and is associated with cognitive deficits. These findings provide crucial theoretical support for the association between cystine and mental health observed in this study, suggesting that cystine may also mediate the influence of glycine, serine, and threonine metabolism on nervous system function [49, 50].

Glycine, threonine, and serine serve as essential or conditionally essential amino acids in humans, playing a dual role in maintaining metabolic balance. They not only participate in nutritional competition and symbiotic regulation within the gut microbiota but also connect to the central nervous system via the gut–brain axis, providing a key metabolic link for microbiota-mediated emotional regulation [51]. Glycine, characterized by its simple molecular structure with a single-hydrogen side chain, can be synthesized from serine through a reversible folate-dependent reaction catalyzed by glycine decarboxylase (GDC, also termed the glycine cleavage system or glycine dehydrogenase) and serine hydroxymethyltransferase (SHMT), both of which facilitate the interconversion between glycine and serine. As a neurotransmitter whose full functional scope remains incompletely understood, glycine occupies a central position in serine and threonine metabolism, is released by specific neurons, and acts as an agonist at N-methyl-D-aspartate receptors (NMDARs), exerting significant influence on neural circuits and neuronal excitability. Moreover, its interaction with the gut microbiota may shape its functional roles within the central nervous system [52, 53]. Serine, a metabolic precursor of glycine, is critical for excitatory glutamatergic neurotransmission; its derivative D-serine, generated under the catalysis of serine racemase, contributes to synaptic plasticity regulation [54, 55]. Threonine (Thr), an essential amino acid that must be obtained from the diet in animals, is degraded by threonine dehydrogenase (TDH) to yield glycine, which in turn regulates protein synthesis by activating mTORC1 in a PI3K/Akt-dependent manner and by inhibiting proteolysis [56, 57]. A key finding of this study is that the core metabolic pathways implicated by gut microbiota-associated metabolites—glycine, threonine, and serine—were independently identified as critical pathways for emotional instability in analyses conducted apart from the microbiota. This result cross-validates the specificity and robustness of the identified signals, demonstrating that the causal chain “gut microbiota *→* metabolites *→* emotional instability” established through MR screening is not a statistical artifact. Instead, it points precisely to a biologically coherent system, supported by independent evidence linking these pathways to emotional regulation. These insights further clarify the specific role of this metabolic axis in emotional instability and help bridge a mechanistic gap in the association between gut microbiota, amino acid metabolism, and emotional dysregulation.

It is noteworthy that the gut microbiota and metabolites identified in this study demonstrate considerable translational potential. Odds ratios (ORs) derived from the multi-step MR framework indicate that gut microbiota such as CAG-485 sp002362485 exert a significant influence on emotional instability as notable risk factors. These findings suggest that the identified microbial and metabolic signatures may not only serve as biomarkers for emotional lability but also offer novel pathways for therapeutic intervention and future research. For example, restoring imbalanced gut microbiota through probiotics, prebiotics, or faecal microbiota transplantation could constitute effective treatment strategies, while screening for corresponding serum metabolites might provide a feasible approach for detecting emotional lability[13]. Concurrently, dietary components play a crucial role in shaping the gut microbiota and modulating host physiology.

Dietary modifications can alter microbial metabolic output, reduce inflammatory markers, and thereby influence mood states [35, 42].

Several limitations of this study warrant consideration. First, although Mendelian randomization reduces confounding and reverse causality, it depends on the validity of instrumental variables and cannot fully rule out weak instrument bias. Second, the gut microbiota GWAS data used were primarily derived from European-ancestry populations, which may limit the generalizability of our findings to other ethnic groups. Finally, while the MR framework helped identify putative gut microbiota-specific mediating pathways, these pathways require further experimental and longitudinal validation.

## Conclusion

In summary, this multi-step Mendelian randomization study provides robust genetic evidence for a causal role of gut microbiota in emotional lability, and reverse MR analyses rule out reverse causality. We identify three gut bacterial taxa with independent causal effects, among which *CAG-485 sp002362485* acts as an emotion lability-specific risk factor. Its pro-risk effect is partially mediated by the glycine, serine and threonine metabolic pathway, with three circulating metabolites identified as key mediators. Our findings delineate a “gut microbiota–amino acid metabolism–emotional regulation” causal axis along the gut-brain axis, and highlight microbial and metabolic signatures as promising diagnostic biomarkers and therapeutic targets for emotional dysregulation. Further validation in diverse ancestral populations and experimental studies are warranted to support clinical translation.

## Data Availability

All genome-wide association study summary statistics used in this Mendelian randomization analysis are publicly available from previously published studies and public repositories. All relevant data generated by the present study, including analysis code and full result tables, are provided in the manuscript and its Supporting Information files.

## Acknowledgements

We acknowledge the FINRISK, FinnGen, and CLSA consortia for providing GWAS summary statistics.

## Supporting information

**S1 Table.** STROBE-MR checklist of recommended items to address in reports of Mendelian randomization studies.

**S2 Table.** *M* ultivariable Mendelian Randomization (MVMR) analysis of the causal effect of gut microbiota (GM) on early Emotion Lability(EL).

**S3 Table.** Results of two-sample MR analysis showing significant associations (P ¡ 0.05) between gut microbiota (GM) and other psychiatric disorders using the IVW method, with MR Egger and weighted median results provided for comparison.

**S4 Table.** MVMR Various Relevant Gut Microbiota Taxa on Bipolar Disorder: A Multivariable Mendelian Randomization (MVMR) Analysis

**S5 Table.** MVMR Various Relevant Gut Microbiota Taxa on PTSD: A Multivariable Mendelian Randomization (MVMR) Analysis

**S6 Table.** Top 20 metabolites mediating the effects of CAG-485 sp002362485 on emotional lability (EL)

